# The association between sarcopenia in the elderly population of China and the risk of falls: a prospective cohort study from CHARLS

**DOI:** 10.64898/2025.12.23.25342898

**Authors:** Xia Yang, Xue Wang, Aiqin Zhao, Haiye Wang, Shan Zheng

## Abstract

**Background:** Sarcopenia is associated with increased fall risk, but its graded relationship with severity and age-specific patterns warrants further investigation in middle-aged and older adults.

**Objective:** To investigate the association between sarcopenia severity and fall risk among adults aged ≥45 years and provide evidence for early intervention.

**Methods:** Utilizing data from the China Health and Retirement Longitudinal Study (CHARLS) database (baseline 2011 to follow-up 2020), 9,608 participants without sarcopenia at baseline were included. Binary logistic regression analyses (univariate, multivariate, and interaction analyses) were employed to assess the association between sarcopenia severity and fall risk. The nonlinear effect of age was examined using restricted cubic splines (RCS curves).

**Results:** The overall fall incidence rate among participants with sarcopenia was 18.88% (1,814/9,608). Fall incidence exhibited a graded increase with sarcopenia severity: non-sarcopenia group: 16.89% (1,108/6,560), possible sarcopenia group: 21.54% (439/2,038), confirmed sarcopenia group: 26.11% (217/831), and severe sarcopenia group: 27.93% (50/179). Multivariate logistic regression revealed a graded increase in fall risk with sarcopenia severity: possible sarcopenia (OR=1.28,CI[1.13-1.45]), confirmed sarcopenia (OR=1.36,CI[1.14-1.63]), and severe sarcopenia (OR=1.44,CI[1.02-2.03]). RCS curves identified a biphasic risk pattern: a modest rise between ages 45-60 and a steep increase after age 70, with 70 years as the inflection point. The subgroup analysis revealed that fall risk was significantly elevated in individuals with sarcopenia who were male, or who had a history of smoking, alcohol consumption, stroke, arthritis, or pain. Key interactions included a 2.96-fold risk in those aged 45-60 with severe sarcopenia (OR=2.96,[1.24-7.06]) and a 4.46-fold risk in those with confirmed sarcopenia and stroke (OR=4.46,[1.45-13.68]).

**Conclusion:** Sarcopenia is an independent risk factor for falls, with risk increasing in a severity-dependent graded manner. Early identification of high-risk individuals and implementation of graded interventions are crucial.

## Introduction

Sarcopenia is an aging syndrome characterized by the loss of muscle mass, muscle strength, and physical function, closely associated with advancing age ^[1]^. Conservative estimates suggest that over 50 million people worldwide are affected by sarcopenia, a number projected to exceed 200 million in the next 40 years ^[2]^. It affects 10-16% of older adults ^[3]^, with prevalence rates of 5%-13% among those over 60 years old, rising to as high as 50% among those over 80 ^[4]^. The prevalence of severe sarcopenia ranges from 2% to 9% ^[5]^. Sarcopenia is strongly associated with multiple adverse outcomes. Its harm extends beyond the degeneration of the muscular system, leading to physical functional decline and disability, increasing the risk of mild cognitive impairment by 1.625 times ^[6]^, hospitalization risk by 40% compared to non-sarcopenic individuals, and cardiovascular disease prevalence up to 18% ^[7]^. As the condition progresses, it exacerbates the risk of falls and fractures ^[8]^, while also increasing mortality risk ^[9]^, profoundly impacting physical health, quality of life, and imposing a significant burden on individuals, families, and socioeconomic systems.

A fall is defined as a sudden, involuntary, unintentional change in position resulting in landing on the same level or a lower level than the starting position ^[10]^. Reported fall incidence rates in China range from 10.3% to 44% ^[11]^, with a rate of 17.42% among the middle-aged and elderly population over 45 years old ^[12]^. The incidence among outpatients and inpatients is 28.8% ^[13]^, and approximately 40% of adverse events in hospitals are falls ^[14]^. Recent studies have found that individuals with declining physical function, low grip strength, and reduced muscle strength (especially in the lower limbs) have a 1.76 times higher fall risk and a 3.06 times higher risk of recurrent falls compared to normal individuals ^[15]^, making it a significant contributing factor to falls in older adults ^[16]^. According to the World Health Organization (WHO), falls are the second leading cause of unintentional injury and death globally, accounting for 0.85% to 1.5% of total global healthcare expenditure. This represents a costly burden for public health systems, constituting approximately 28-35% of related costs ^[17]^. While sarcopenia was traditionally considered a disease of old age (>65 years), increasing evidence suggests that age-related decline in muscle mass and function, and the consequent increased fall risk, actually begin to accelerate during middle age (around 45 years). Recent research on the association between sarcopenia and fall risk is limited and primarily focused on the elderly. Therefore, this study aims to utilize data from the China Health and Retirement Longitudinal Study (CHARLS) in a cohort study design to explore the association between sarcopenia and fall risk among middle-aged and older adults aged 45 and above. This aims to raise awareness about the early identification of sarcopenia, provide evidence for fall prevention, and ultimately promote active and healthy aging to improve the quality of life for this population.

## 1 Research subjects and methods

### 1.1 Research Subjects

CHARLS is a nationally representative longitudinal survey employing a multi-stage cluster sampling method to select Chinese residents aged 45 and older. It covers 150 counties and 450 villages across 28 provinces (autonomous regions, and municipalities), ensuring comprehensive and representative data.

This study extracted data from the CHARLS database baseline (2011) and follow-up waves (2013, 2020), constructing a cohort database excluding individuals with sarcopenia at baseline. Ultimately, 9608 individuals aged 45 and above were included. When constructing the baseline database, since the 2011 and 2013 physical examination data were consistent, duplicate records were removed, retaining only individuals with physical examination data in 2011. Multiple imputation was used for important missing variables. Inclusion criteria: (1) Aged 45 years or older at baseline survey. (2) Participants who completed grip strength and the 5-time chair stand test measurements during the baseline survey. Exclusion criteria: (1) Aged less than 45 years. (2) Missing sociodemographic data, sarcopenia diagnostic data, or follow-up data. (3) Individuals already diagnosed with sarcopenia. The screening flowchart is shown in **Figure 1**.

**Figure 1.**
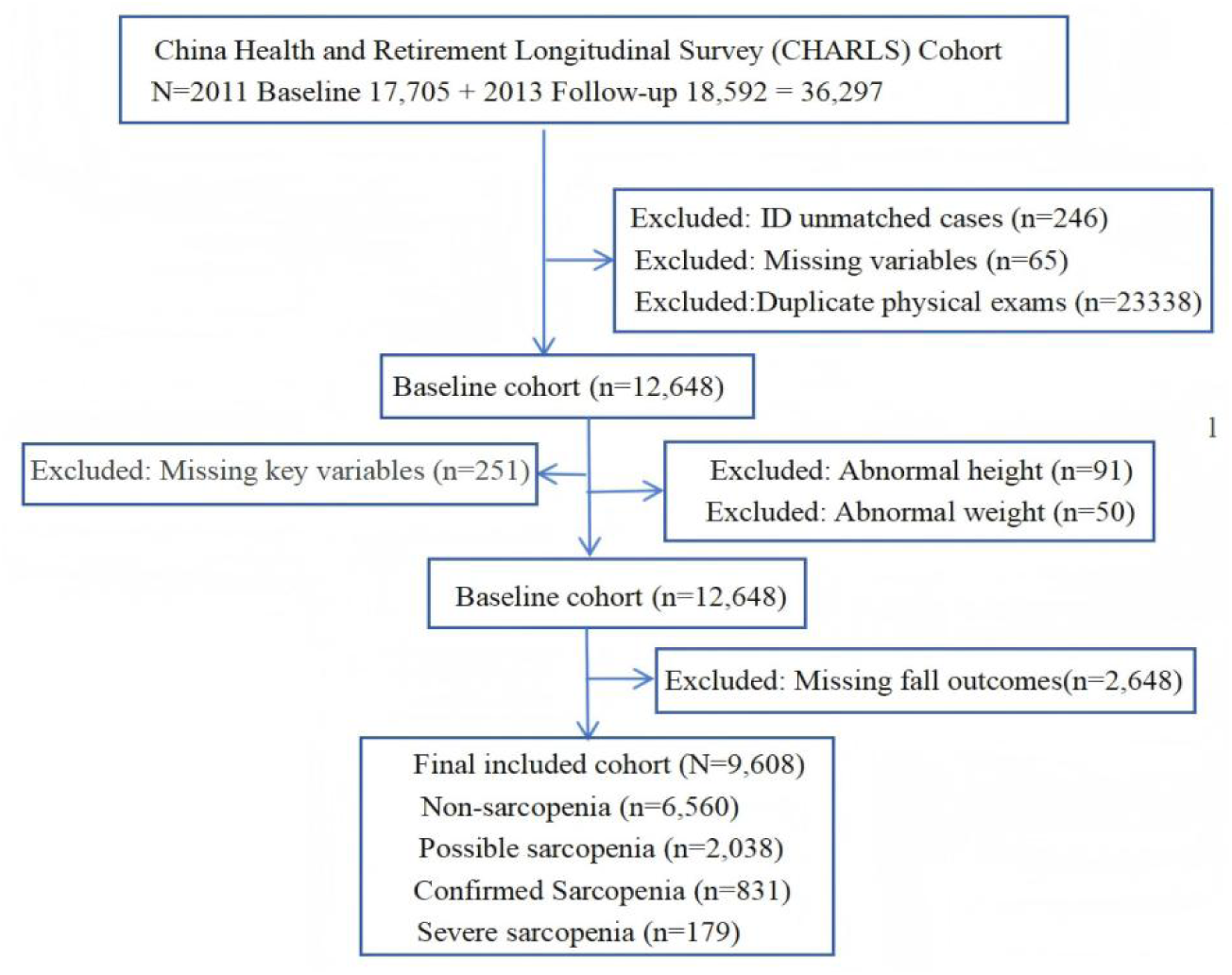
Flowchart of Sarcopenia-Fall Cohort Population Screening.

### 1.2 Ethical Considerations

This study was reviewed and approved by the Ethics Committee of Peking University (Approval No. IRB00001052-11015), and written informed consent was obtained from all participants.

### 1.3 Research Methods

#### 1.3.1 Baseline Survey

A cohort database of middle-aged and older adults was established, with individuals newly enrolled in 2011 and aged ≥45 years without sarcopenia as the baseline population. Survey content included: (1) Demographic factors (gender, age, residence); (2) Health behaviors (smoking, alcohol consumption); (3) Chronic diseases (vision and hearing problems, diabetes, hypertension, hyperlipidemia, arthritis, hip fracture, pain). For demographic factors: gender coded as 1=Male, 2=Female; residence coded as 1=Urban, 2=Rural. For lifestyle factors: smoking and alcohol consumption both coded as 1=Yes, 2=No. For chronic diseases: coded as 1=Yes (diagnosed), 2=No (not diagnosed).

#### 1.3.2 Follow-up Survey

Participants from the 2011 baseline were followed up in 2020. The primary outcome was the occurrence of falls. Determination method: In the 2020 follow-up data, occurrence of a fall was coded as 1=Yes, non-occurrence as 2=No.

### 1.4 Definitions and Diagnostic Criteria

Sarcopenia was diagnosed according to the 2019 Asian Working Group for Sarcopenia (AWGS 2019) consensus criteria :(1) Non-sarcopenia: Normal muscle mass, muscle strength, and physical function.(2) Possible sarcopenia: Positive screening result plus low muscle strength or low physical function.(3)confirmed Sarcopenia: Low muscle mass plus low muscle strength or low physical function.(4) Severe sarcopenia: Low muscle mass plus low muscle strength plus low physical function^[18]^.Sarcopenia assessment involved three dimensions:Muscle Strength: Assessed by handgrip strength, using the maximum value from left and right hand measurements. Low muscle strength was defined as <28 kg for men and <18 kg for women (measured using Yuejian WL-1000 dynamometer).Physical Function: Assessed by any of the following indicating low physical performance ^[19]^: Short Physical Performance Battery (SPPB) total score ≤9; usual gait speed (GS) <1.0 m/s; 5-time chair stand test time >12 seconds.Muscle Mass: Measured by dual-energy X-ray absorptiometry (DXA) or bioelectrical impedance analysis (BIA). Appendicular skeletal muscle mass (ASM) was calculated. Appendicular skeletal muscle mass index (ASMI) = ASM (kg) / height² (m²). AWGS 2019 defines low muscle mass as ASMI <7.0 kg/m² for men and <5.4 kg/m² (BIA) or <5.7 kg/m² (DXA) for women. AWGS 2019 recommends BIA for muscle mass measurement ^[20]^. The formula used for ASM (presumably from BIA) was:

ASM = 0.193 × weight (kg) + 0.107 × height (cm) - 4.157 × gender - 0.037 × age (years) - 2.631.

### 1.5 Statistical Methods

Statistical analysis was performed using Stata 18.0 software. Frequency (percentage) was used to describe the distribution of exposure factors. Binary logistic regression was employed, incorporating variables significant in univariate analysis. Subgroup analysis and RCS modeling were conducted.(*P-value* < 0.05 was considered statistically significant.)

## 2 Results

### 2.1 Baseline Characteristics and Fall Incidence by Sarcopenia Status

A total of 9608 middle-aged and older adults ≥45 years were included at baseline. The overall fall incidence rate among those who developed sarcopenia was 18.88% (1814/9608), increasing progressively with sarcopenia severity: non-sarcopenia group 16.89% (1108/6560), possible sarcopenia group 21.54% (439/2038), confirmed sarcopenia group 26.11% (217/831), severe sarcopenia group 27.93% (50/179). Differences between all sarcopenia groups regarding age, gender, residence, smoking, alcohol consumption, and chronic diseases were statistically significant (*P*<0.05). Baseline population characteristics are detailed in **Table 1**. Fall incidence rates are shown in **Table 2**.

**Table 1.**
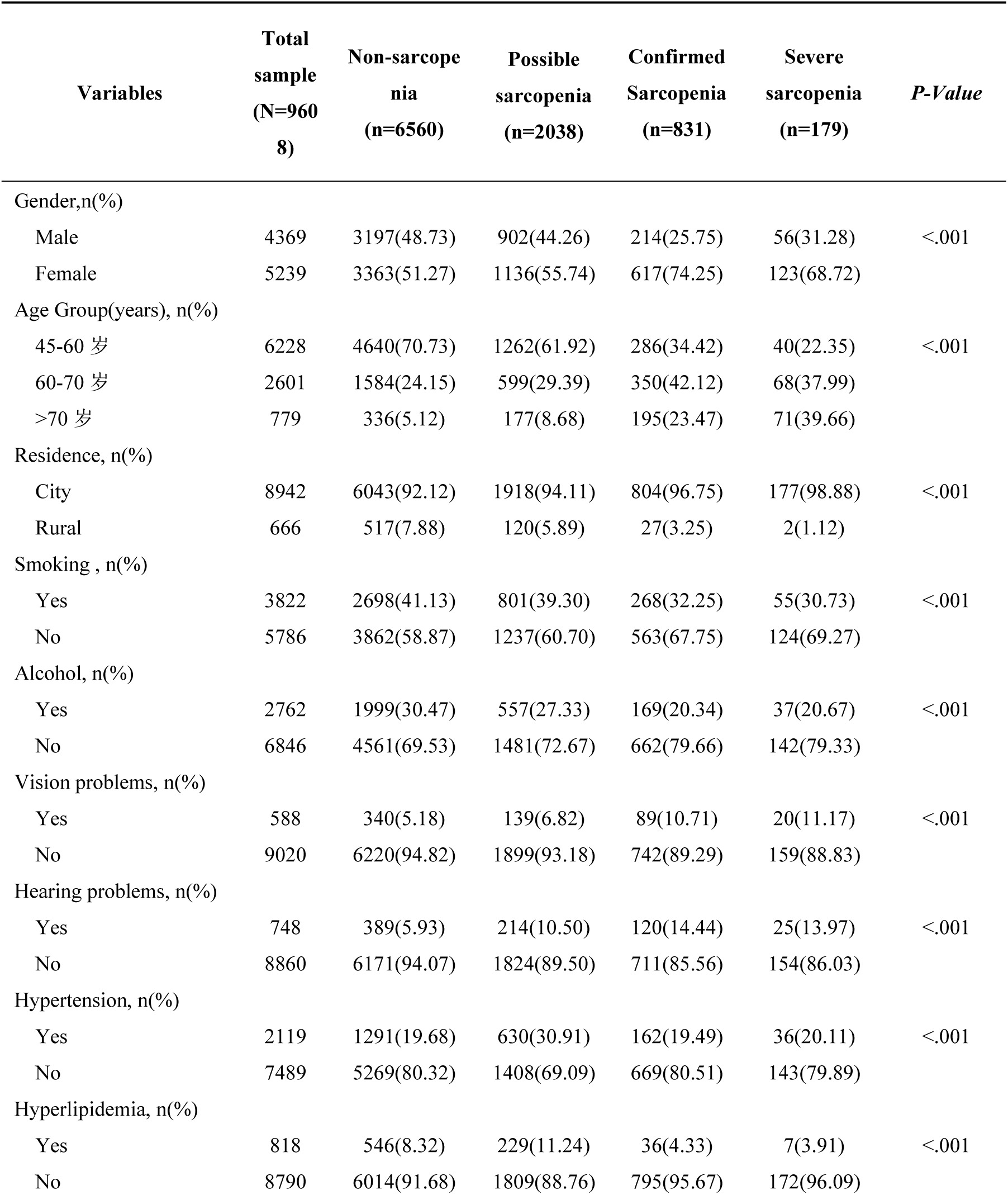
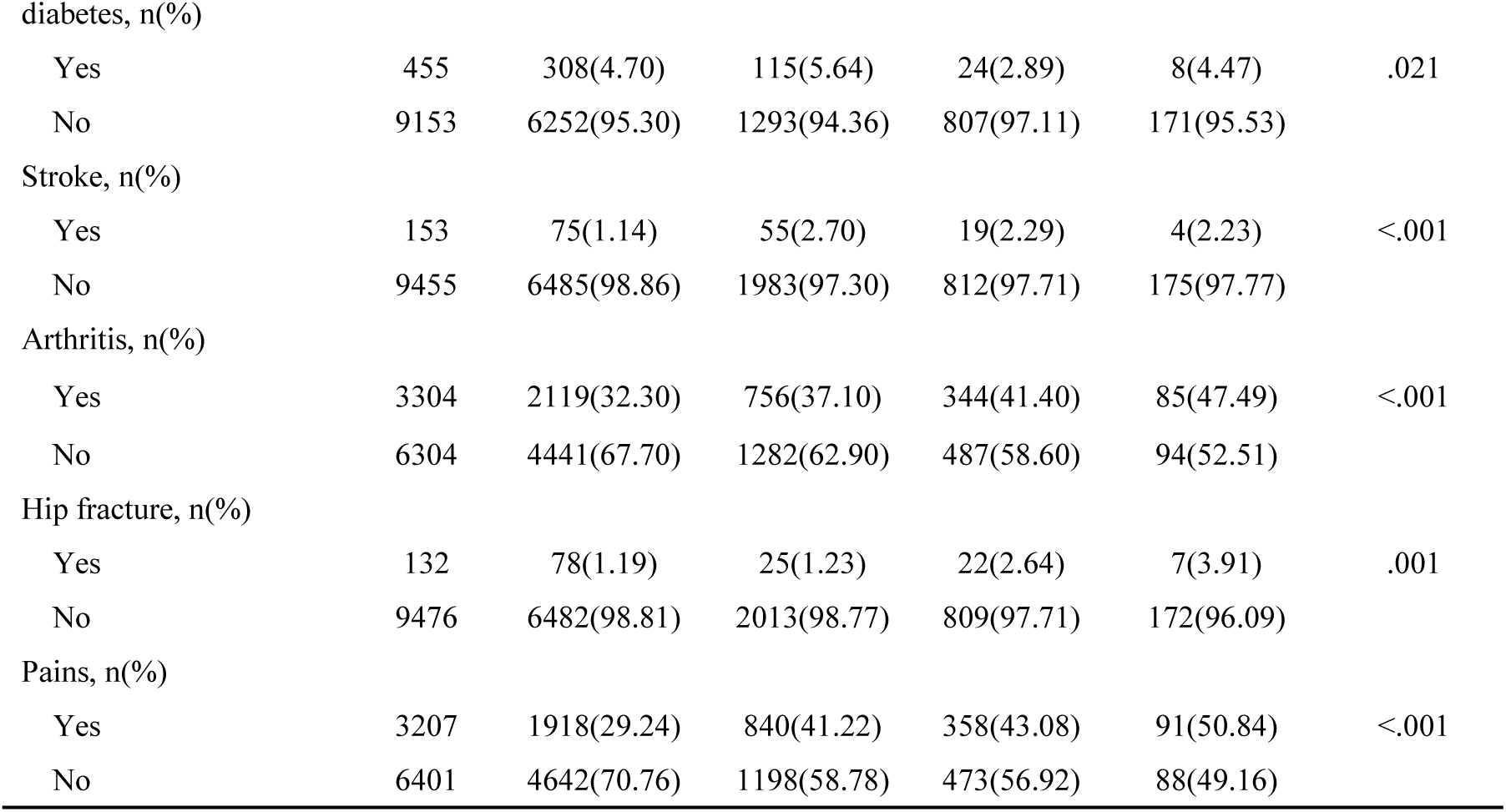
Baseline Characteristics of the Sarcopenia Cohort Population Aged ≥45 Years.

| Variables | Total sample<br>(N=9608) | Non-sarcope<br>nia<br>(n=6560) | Possible<br>sarcopenia<br>(n=2038) | Confirmed<br>Sarcopenia<br>(n=831) | Severe<br>sarcopenia<br>(n=179) | *P-Value* |
| --- | --- | --- | --- | --- | --- | --- |
| Gender,n(%) |  |  |  |  |  |  |
| Male | 4369 | 3197(48.73) | 902(44.26) | 214(25.75) | 56(31.28) | <.001 |
| Female | 5239 | 3363(51.27) | 1136(55.74) | 617(74.25) | 123(68.72) |  |
| Age Group(years), n(%) |  |  |  |  |  |  |
| 45-60 岁 | 6228 | 4640(70.73) | 1262(61.92) | 286(34.42) | 40(22.35) | <.001 |
| 60-70 岁 | 2601 | 1584(24.15) | 599(29.39) | 350(42.12) | 68(37.99) |  |
| >70 岁 | 779 | 336(5.12) | 177(8.68) | 195(23.47) | 71(39.66) |  |
| Residence, n(%) |  |  |  |  |  |  |
| City | 8942 | 6043(92.12) | 1918(94.11) | 804(96.75) | 177(98.88) | <.001 |
| Rural | 666 | 517(7.88) | 120(5.89) | 27(3.25) | 2(1.12) |  |
| Smoking , n(%) |  |  |  |  |  |  |
| Yes | 3822 | 2698(41.13) | 801(39.30) | 268(32.25) | 55(30.73) | <.001 |
| No | 5786 | 3862(58.87) | 1237(60.70) | 563(67.75) | 124(69.27) |  |
| Alcohol, n(%) |  |  |  |  |  |  |
| Yes | 2762 | 1999(30.47) | 557(27.33) | 169(20.34) | 37(20.67) | <.001 |
| No | 6846 | 4561(69.53) | 1481(72.67) | 662(79.66) | 142(79.33) |  |
| Vision problems, n(%) |  |  |  |  |  |  |
| Yes | 588 | 340(5.18) | 139(6.82) | 89(10.71) | 20(11.17) | <.001 |
| No | 9020 | 6220(94.82) | 1899(93.18) | 742(89.29) | 159(88.83) |  |
| Hearing problems, n(%) |  |  |  |  |  |  |
| Yes | 748 | 389(5.93) | 214(10.50) | 120(14.44) | 25(13.97) | <.001 |
| No | 8860 | 6171(94.07) | 1824(89.50) | 711(85.56) | 154(86.03) |  |
| Hypertension, n(%) |  |  |  |  |  |  |
| Yes | 2119 | 1291(19.68) | 630(30.91) | 162(19.49) | 36(20.11) | <.001 |
| No | 7489 | 5269(80.32) | 1408(69.09) | 669(80.51) | 143(79.89) |  |
| Hyperlipidemia, n(%) |  |  |  |  |  |  |
| Yes | 818 | 546(8.32) | 229(11.24) | 36(4.33) | 7(3.91) | <.001 |
| No | 8790 | 6014(91.68) | 1809(88.76) | 795(95.67) | 172(96.09) |  |
| diabetes, n(%) |  |  |  |  |  |  |
| Yes | 455 | 308(4.70) | 115(5.64) | 24(2.89) | 8(4.47) | .021 |
| No | 9153 | 6252(95.30) | 1293(94.36) | 807(97.11) | 171(95.53) |  |
| Stroke, n(%) |  |  |  |  |  |  |
| Yes | 153 | 75(1.14) | 55(2.70) | 19(2.29) | 4(2.23) | <.001 |
| No | 9455 | 6485(98.86) | 1983(97.30) | 812(97.71) | 175(97.77) |  |
| Arthritis, n(%) |  |  |  |  |  |  |
| Yes | 3304 | 2119(32.30) | 756(37.10) | 344(41.40) | 85(47.49) | <.001 |
| No | 6304 | 4441(67.70) | 1282(62.90) | 487(58.60) | 94(52.51) |  |
| Hip fracture, n(%) |  |  |  |  |  |  |
| Yes | 132 | 78(1.19) | 25(1.23) | 22(2.64) | 7(3.91) | .001 |
| No | 9476 | 6482(98.81) | 2013(98.77) | 809(97.71) | 172(96.09) |  |
| Pains, n(%) |  |  |  |  |  |  |
| Yes | 3207 | 1918(29.24) | 840(41.22) | 358(43.08) | 91(50.84) | <.001 |
| No | 6401 | 4642(70.76) | 1198(58.78) | 473(56.92) | 88(49.16) |  |

**Table 2.** Fall Incidence Rate.

| Sarcopenia Category | Total<br>(N=9608) | Fall case<br>(n=1814) | Fall Incidence<br>(%) | Fall Incidence<br>95%CI |
| --- | --- | --- | --- | --- |
| Non-sarcopenia | 6560 | 1108 | 16.89 | 16.00-17.81 |
| Possible Sarcopenia | 2038 | 439 | 21.54 | 19.81-23.38 |
| Confirmed Sarcopenia | 831 | 217 | 26.11 | 23.24-29.20 |
| Severe sarcopenia | 179 | 50 | 27.93 | 21.87-34.91 |
| Total | 9608 | 1814 | 18.88 | 18.10-19.66 |

### 2.2 Logistic Regression Results for Fall Risk

#### 2.2.1 Univariate Logistic Results

Variables included in the univariate analysis (demographics: gender, age, residence; health behaviors: smoking, alcohol; chronic diseases: vision problems, hearing problems, hypertension, diabetes, hyperlipidemia, arthritis, hip fracture, pain) were all statistically significant (*P*<0.05). Results of univariate logistic regression for fall risk factors are detailed in **Table 3**.

**Table 3.**
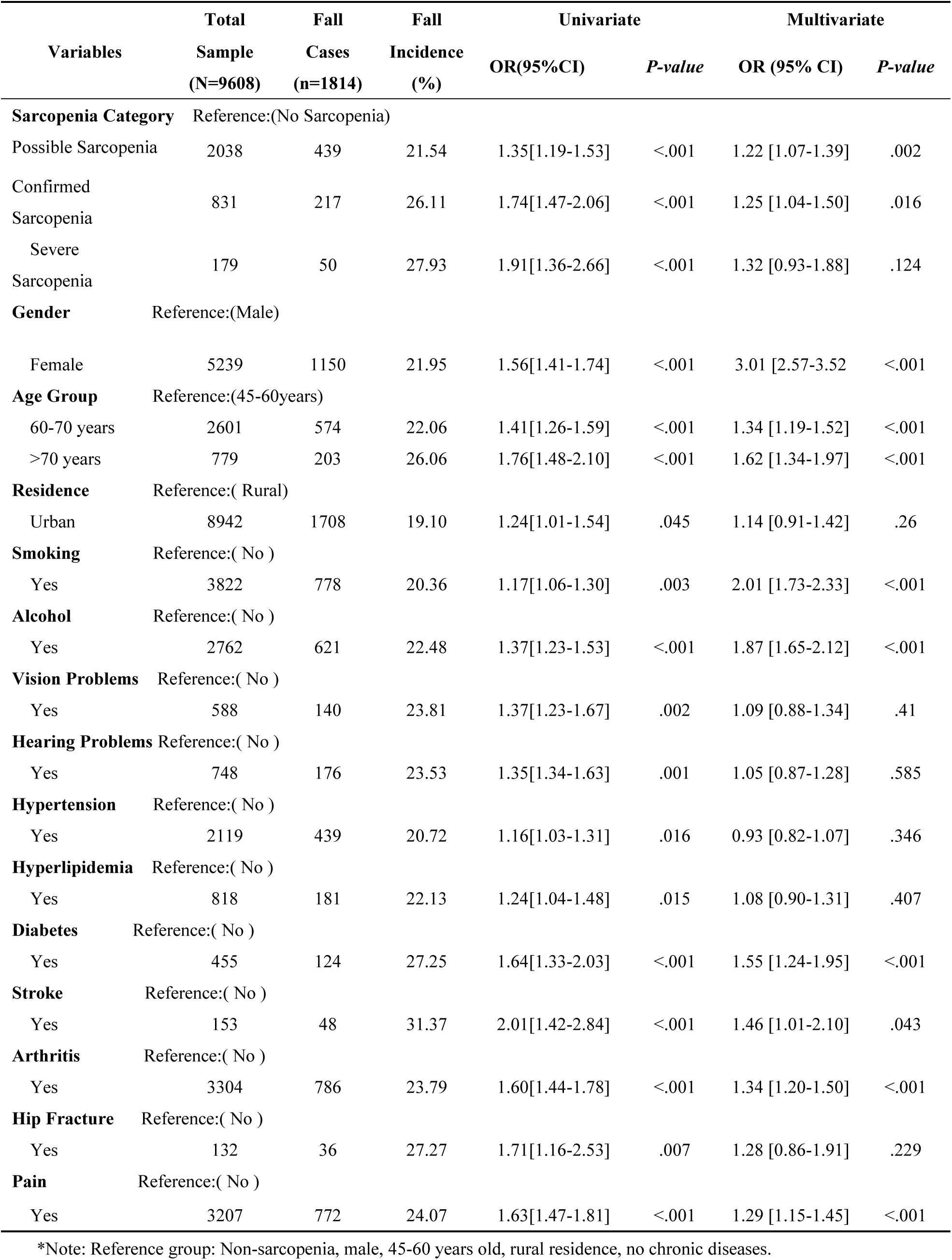
Univariate and Multivariate Logistic Regression Results for Fall Risk

#### 2.2.2 Multivariate Logistic Results

Variables statistically significant in the univariate analysis were included in the multivariate regression. In Model 1 and Model 2, the risk of falls increased significantly across sarcopenia categories (*P*<0.05) (Model 1: Possible: OR=1.28, 95% CI [1.13-1.45]; Confirmed : OR=1.36, 95% CI [1.14-1.63]; Severe: OR=1.44, 95% CI [1.02-2.03]). In Model 3 (fully adjusted), significant increases in fall risk were observed for the possible sarcopenia group, Confirmed sarcopenia group, females, age groups 60-70 years and >70 years, smoking, alcohol consumption, diabetes, stroke, arthritis, and pains (*P*<0.05). Results are detailed in **Table 3** and summarized for sarcopenia categories in **Table 4**.

**Table 4.** Multivariate Logistic Regression Results for Sarcopenia and Fall Risk

| Variables | Model 1 |  | Model 2 |  | Model 3 |  |
| --- | --- | --- | --- | --- | --- | --- |
| Sarcopenia Category | OR[95%CI] | *P-value* | OR[95%CI] | *P-value* | OR[95%CI] | *P-value* |
| Possible Sarcopenia | 1.28[1.13-1.45] | <.001 | 1.29[1.14-1.46] | <.001 | 1.21 [1.07-1.37] | .003 |
| Confirmed Sarcopenia | 1.36[1.14-1.63] | .001 | 1.37[1.15-1.63] | .001 | 1.31 [1.10-1.56] | .003 |
| Severe Sarcopenia | 1.44[1.02-2.03] | .038 | 1.44[1.02-2.03] | .038 | 1.31 [0.92-1.85] | .132 |

\*Note: Reference group: Non-sarcopenia. Model 1: Unadjusted. Model 2: Adjusted for gender, age, smoking, alcohol consumption. Model 3: Adjusted for gender, age,, smoking, alcohol consumption, diabetes, stroke, arthritis, pain.

### 2.3 Results of the Subgroup Analysis on Fall Risk in Sarcopenia

Subgroup analyses based on sex, age group, smoking, alcohol consumption, diabetes, stroke, arthritis, and pain revealed the following: (1) Sex differences: Sarcopenia was associated with a significantly increased risk of falls in males across all subgroups (with an OR = 2.16 in the severe sarcopenia group). In females, a significantly increased risk was observed only in the confirmed sarcopenia and severe sarcopenia groups. (2) Age stratification: In the 45–60 years age group, all sarcopenia subgroups showed an increased risk of falls (with an OR = 3.35 in the severe sarcopenia group). In the 60–70 years age group, only the confirmed sarcopenia group exhibited a significantly increased risk (OR = 1.44). No significant association was observed in the >70 years age group. (3) Behavioral and disease factors: Compared with controls, a significantly higher risk of falls was found among individuals with sarcopenia who smoked, consumed alcohol, had arthritis (except in the severe sarcopenia subgroup), had stroke (except in the possible sarcopenia subgroup), or experienced pain. However, no significantly increased risk was observed among those with sarcopenia and diabetes. Detailed results are presented in **Figure 2**.

**Figure 2.**
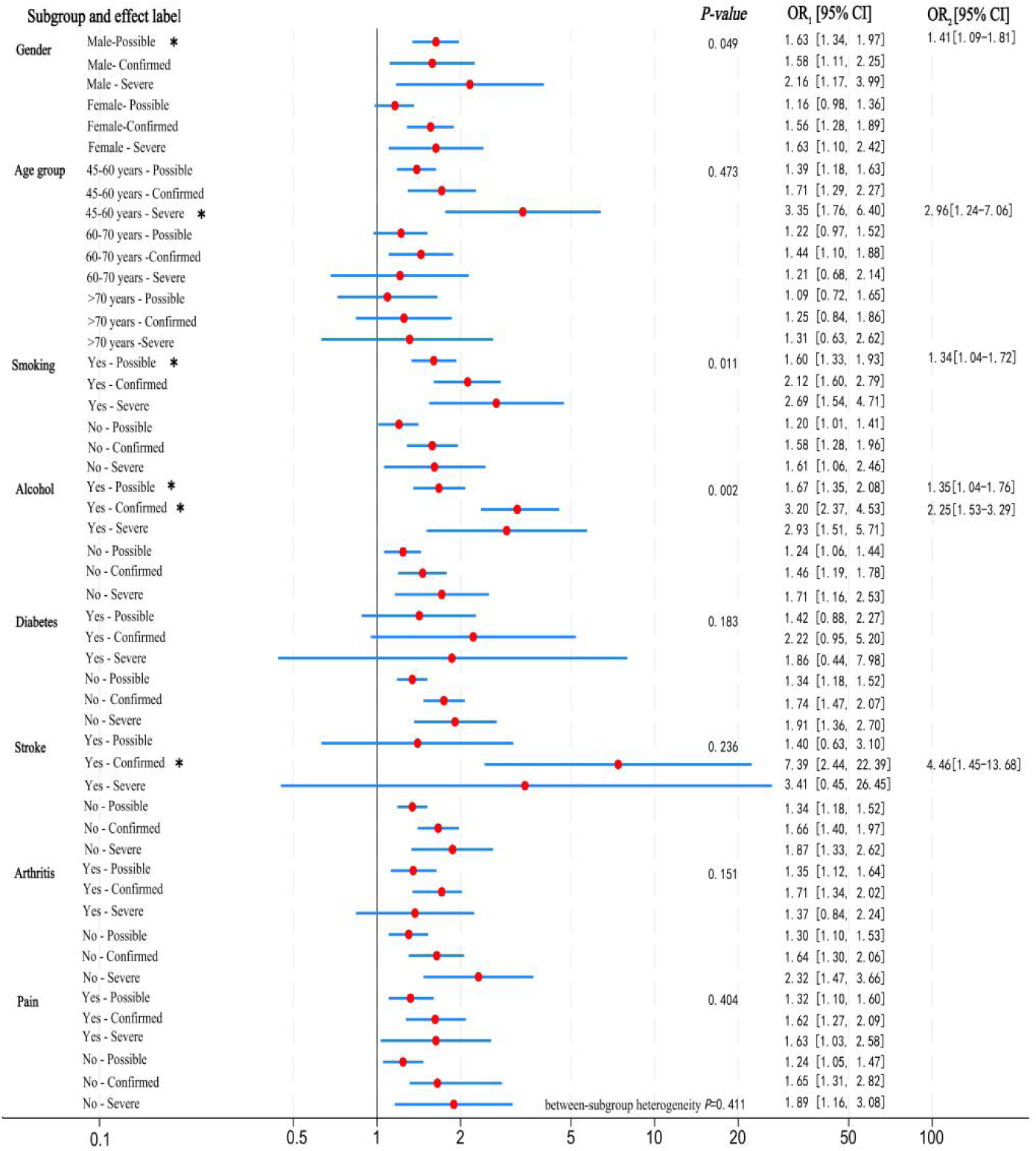
Forest Plot of Sarcopenia and Fall Risk. Note: Legend for Figure 2:OR_1_: Odds ratio of the subgroup variable.OR_2_: Odds ratio of the interaction term.**\***:Interaction term.

### 2.4 Sarcopenia Interaction Results

A significantly increased risk of falls was observed in the following subgroups: males with possible sarcopenia had a 41% higher risk (OR = 1.41, 95% CI [1.09-1.81]); individuals with possible sarcopenia who smoked exhibited a 1.34-fold increased risk (OR = 1.34, 95% CI [1.04-1.72]); among those who consumed alcohol, the possible sarcopenia and confirmed sarcopenia groups showed a 1.35-fold (OR = 1.35, 95% CI [1.04-1.76]) and a 2.25-fold (OR = 2.25, 95% CI [1.53-3.29]) increase in risk, respectively; and patients with confirmed sarcopenia and a history of stroke had a 4.46-fold higher risk (OR = 4.46, 95% CI [1.45-13.68]) (*P* < 0.05). Detailed results of the interaction analysis for sarcopenia are presented in **Table 5**.

**Table 5.** Sarcopenia Interaction Results

| Variables | OR | [95% CI] | P-value | Variables | OR | [95% CI] | P-value |
| --- | --- | --- | --- | --- | --- | --- | --- |
| **Sarcopenia** |  |  |  | **Sarcopenia** |  |  |  |
| **Category#Gender** |  |  | <.001 | **Category#Age Group** |  |  | <.001 |
| **Possible#Male** | **1.41** | **[1.09-1.81]** | **.008** | Possible#45-60years | 1.28 | [0.81-1.99] | .286 |
| Confirmed#Male | 1.02 | [0.68-1.52] | .939 | Confirmed#45-60years | 1.37 | [0.84-2.24] | .207 |
| Severe#Mal | 1.32 | [0.63-2.75] | .448 | **Severe #45-60years** | **2.96** | **[1.24-7.06]** | **.014** |
| Possible#Female | 0.71 | [0.55-0.91] | .008 | Possible#60-70years | 0.88 | [0.67-1.16] | .352 |
| Confirmed#Female | 0.98 | [0.66-1.47] | .939 | Confirmed#60-70years | 0.84 | [0.57-1.24] | .385 |
| Severe#Female | 0.75 | [0.36-1.57] | .448 | Severe#60-70years | 0.40 | [0.15-0.85] | .020 |
| **SarcopeniaCategory** |  |  |  | Possible#>70years | 0.78 | [0.50-1.23] | .286 |
| **#Smoking** |  |  | <.001 | Confirmed#>70years | 0.73 | [0.45-1.19] | .207 |
| **Possible#Smoking** | **1.34** | **[1.04-1.72]** | **.022** | Severe#>70years | 0.34 | [0.14-0.80] | .014 |
| Confirmed#Smoking | 1.34 | [0.94-1.89] | .104 |  |  |  |  |
| Severe#Smoking | 1.66 | [0.83-3.35] | .153 |  |  |  |  |
| **Sarcopenia** |  |  |  | **Sarcopenia** |  |  |  |
| **Category#Alcohol** |  |  | <.001 | **Category#Stroke** |  |  |  |
| **Possible#Alcohol** | **1.35** | **[1.04-1.76]** | **.025** | Possible#Stroke | 1.04 | [0.47-2.33] | .916 |
| **Confirmed#Alcohol** | **2.25** | **[1.53-3.29]** | **<.001** | **Confirmed#Stroke** | **4.46** | **[1.45-13.68]** | **.009** |
| Severe#Alcohol | 1.71 | [0.79-3.70] | .172 | Severe#Stroke | 1.83 | [0.23-14.34] | .567 |

Note: The figure shows interactions of sarcopenia category with sex, age group, smoking, alcohol consumption, and stroke.

### 2.5 RCS curve for sarcopenia fall risk

RCS curves were plotted after adjusting for gender, residence, diabetes, stroke, arthritis, and pain. Results demonstrated:(1) Overall Trend: Sarcopenia-related fall risk exhibited a dual-rising pattern: a gradual increase between ages 45 and 60, followed by a steep rise after age 70, with age 70 identified as a significant inflection point.(2) Differences by Sarcopenia Severity:Possible Sarcopenia: Fall risk increased progressively with advancing age without significant inflection points.Confirmed Sarcopenia: Prevalence rose significantly in the age 45-60, stabilized between ages 60 and 70, and surged sharply after age 70.Severe Sarcopenia: Risk escalated rapidly between ages 45 and 55, unexpectedly declined from age 55 to 70, then rose sharply again after age 70, indicating a triphasic age-dependent trajectory.(RCS curve results shown in **Figure 3**.)

**Figure 3.**
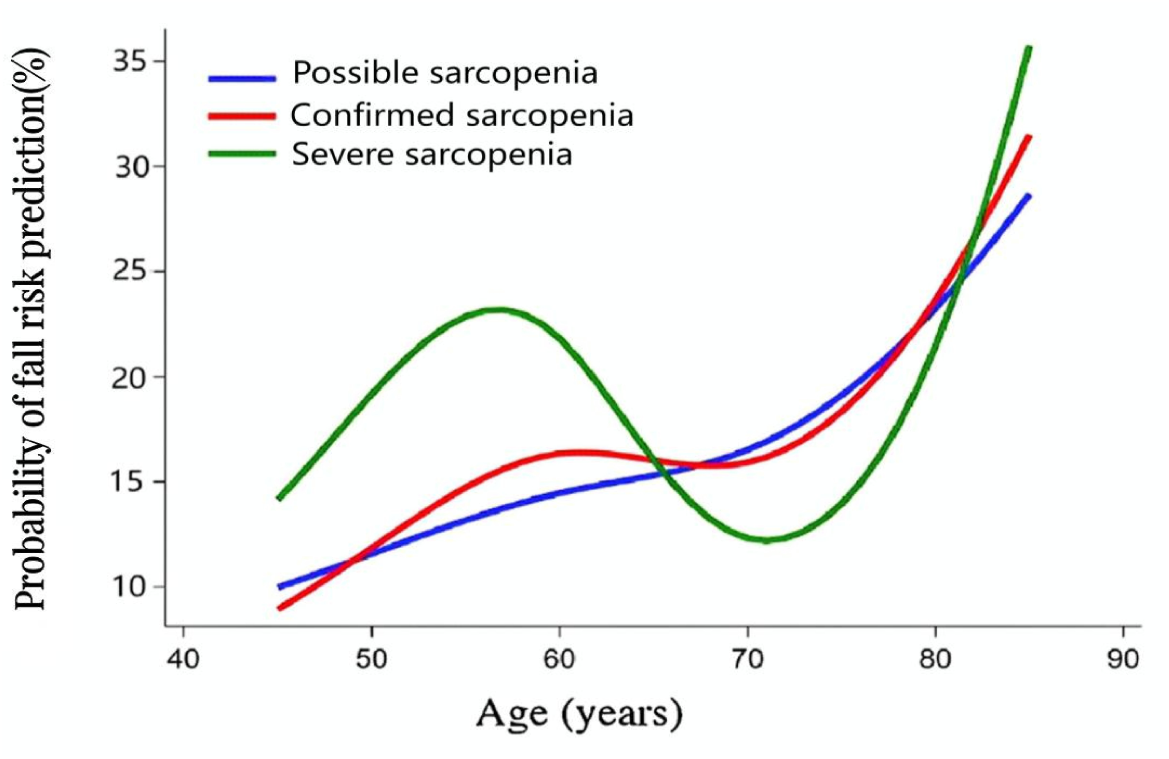
RCS Curves of Sarcopenia and Fall Risk.

## 3 Discussion

This study confirms a dose-response relationship between sarcopenia severity and fall risk. As sarcopenia progressed from possible, to confirmed, and finally to severe, fall risk increased in a graded manner (by 28%, 36%, and 44%, respectively), and fall incidence similarly rose (21.54%, 26.11%, and 27.93%, respectively). These findings align with previous research^[21, 22]^. The core mechanism lies in the progressive functional decline of the muscle-neuro-skeletal system^[23]^, with type I and II muscle fiber atrophy being particularly critical^[24]^. Additionally, aging-associated inflammation, oxidative stress, mitochondrial dysfunction, and apoptosis contribute^[25]^. Specific manifestations include:(1) Structural Degeneration: The proportion of fast-twitch muscle fibers significantly decreases, from approximately 60% in young men to about 30% in 80-year-old men^[26]^. Accelerated muscle fiber atrophy leads to a marked reduction in muscle strength, with a decline of 16.6% to 40.9% observed in individuals over 40 years old compared to those under 40^[27]^.(2) Neuromotor dDecompensation: With aging, the loss of motor neurons (particularly α-motor neurons) and degradation of muscle contractility cannot be fully compensated for by the reinnervation of muscle fibers by remaining motor neurons, resulting in a continuous decline in muscle mass and function^[28]^. Age-related neural impairment has been confirmed to be closely associated with sarcopenia in the elderly^[29]^.(3) Activation of Molecular Pathways: Mitochondrial dysfunction within skeletal muscle cells is considered a primary driver of sarcopenia^[30, 31]^. These processes form a vicious cycle: the more severe the sarcopenia, the greater the skeletal muscle force required to maintain posture, leading to accelerated failure of fast-twitch fibers and a multiplied fall risk.

This study further revealed age-specific fall risk patterns. Restricted cubic spline (RCS) curves indicated two distinct periods of significant risk increase: 45-55 years and after 70 years. Within these age ranges, the curves exhibited a pronounced upward trend, signifying a steep rise in fall risk with advancing age.Age 45-55 Stage: Risk is primarily dominated by the functional collapse of fast-twitch fibers. Muscle mass and strength loss accelerate after age 50, with muscle mass declining by approximately 3-8% per decade, and the rate of loss further increasing after the sixth decade^[32]^.After Age 70: Age-related muscle weakness mainly stems from impaired force-generating capacity within individual muscle fibers^[33]^. Elevated levels of interleukin-6 (IL-6), tumor necrosis factor-alpha (TNF-α), and C-reactive protein (CRP) during this stage are significantly associated with a 2- to 3-fold increased risk of muscle strength loss exceeding 40% in the elderly^[34]^.Implications for intervention strategy: The age of 70 serves as a critical inflection point, suggesting the need for staged interventions. Focus before age 70 should be on muscle and neural protection; after age 70, comprehensive interventions centered on anti-inflammation, nutritional support, and disease prevention should be initiated. High-intensity resistance training (HIT-RT) combined with protein, vitamin D, and calcium supplementation has been proven safe and effective for ameliorating sarcopenia in the elderly^[35]^.Furthermore, the sarcopenia-related fall risk curve shows a gradual upward trend starting at age 45, indicating increasing risk with age,This aligns with prior research suggesting individuals with “possible sarcopenia” have a 34% increased fall risk^[36]^. A unique finding within the curve is the “risk decline” observed among individuals with severe sarcopenia aged 55-70. This may reflect the influence of competing risks from comorbidities or behavioral adaptations (e.g., increased bed rest). Studies show that muscle strength declines much faster than muscle atrophy during the first two weeks of bed rest, with fall incidence decreasing to 2.4 by day 14^[37]^. This stage may represent a critical window for intervention to reverse risk. The relatively flat curve between ages 60 and 70 presents an opportunity for interventions to block the abrupt surge in risk after age 70. This study also confirmed that individuals with impaired vision or hearing, diabetes, hyperlipidemia, arthritis, or bodily pain have a significantly higher fall risk, consistent with previous findings^[38]^.

Interaction and subgroup analyses from this study revealed differences in fall risk among the sarcopenic population based on demographic and behavioral/disease factors:(1)Gender Differences: Males with sarcopenia exhibited higher fall risk (OR=2.16 in the severe sarcopenia group), consistent with previous conclusions^[39]^. Male aging is accompanied by significant reductions in muscle and bone mass, with age-related declines in androgen and growth hormone secretion likely playing important roles^[40]^. The age-dependent increase in sarcopenia is more pronounced in males, characterized by faster declines in fast-twitch fiber strength and more significant atrophy^[41]^, with peak power output declining by 50%-55% with age^[42]^. In contrast, the increased fall risk among females with sarcopenia primarily becomes apparent at the stage of confirmed sarcopenia. Females maintain relatively better muscle function during aging^[43]^, potentially due to estradiol stabilizing satellite cell numbers by reducing inflammatory status, thereby partially counteracting the adverse effects of sarcopenia^[44]^. However, dysregulated satellite cell apoptosis caused by sex hormone imbalance remains a potenital trigger for sarcopenia in females^[45]^.(2)Urban-Rural Differences: Rural residents with sarcopenia had significantly higher fall risk than their urban counterparts (OR=6.39 for severe sarcopenia group), likely related to environmental factors, nutritional status, and workload.(3)Behavioral Factors: Smokers and alcohol consumers with sarcopenia had over twice the fall risk of controls, particularly within the confirmed sarcopenia and severe sarcopenia groups. This may be attributed to delayed muscle injury repair, compromised muscle endurance, and impaired vestibular sensory function leading to decreased dynamic balance following smoking and alcohol consumption. Previous research also confirms smoking and alcohol consumption as significant risk factors for falls^[46]^.(4) Comorbidity impact: Subgroup analyses indicated that individuals with confirmed or severe sarcopenia and a history of stroke exhibited 3- to 7-fold higher fall risk. Although stroke itself may cause sensory impairments and balance dysfunction, the small sample size in this subgroup likely resulted in wide 95% confidence intervals (CIs), suggesting imprecise estimates.

Addressing sarcopenia-related fall risk necessitates the construction of a multi-dimensional prevention and control system:(1)In lifestyle interventions, reducing sedentary behavior is as crucial as increasing physical activity. On the one hand, sedentary behavior is a significant risk factor for muscle loss. Evidence indicates that the specific force of fast-twitch muscle fibers can be reduced by up to 31% in sedentary older men ^[33]^.On the other hand, substituting sedentary behavior with active exercise can effectively counteract this risk. Research demonstrates that replacing sedentariness with 30 minutes of moderate-to-vigorous physical activity (MVPA) daily significantly lowers the incidence of sarcopenia ^[47]^.Optimize Nutrition: Avoid excessive intake of processed foods and pro-inflammatory components. Vitamin D insufficiency or deficiency is linked to muscle fiber atrophy,chronic musculoskeletal pain, sarcopenia, and increased fall risk^[48]^. Adequate nutritional intake, particularly protein, is a modifiable factor for sarcopenia and foundational for successful intervention^[49]^. Currently, resistance training (RT) combined with amino acid supplementation is considered the gold standard for preventing sarcopenia^[50]^. This combined physical exercise and nutritional approach is a primary prevention strategy supported by current research^[51]^.Manage Metabolism and Stress: Chronic stress, depression, and obesity exacerbate inflammatory status and muscle catabolism, impairing neuromuscular function^[52]^. Insulin resistance, hyperglycemia, and type 2 diabetes can induce mitochondrial dysfunction, impaired oxidative metabolism, and energy utilization disturbances, leading to sarcopenia^[53]^. Regular monitoring of blood glucose, blood lipids, and body composition is recommended for individuals over 45 years to enable early identification of sarcopenia risk factors such as high-density Lipoprotein and homocysteine ^[54]^.(2)Exercise Management:Emphasizing rapid force and power training is crucial for maintaining fast-twitch fiber function and power output, which is vital for rapid reaction movements and fall prevention^[55]^.In elderly sarcopenic patients, resistance training (RT) and mixed training (MT - combining RT with balance, endurance, or aerobic exercise) effectively improve knee extension strength (KES) and gait speed (GS)^[56]^.Maintaining appropriate frequency and intensity of resistance training can increase muscle mass (particularly type II muscle fibers) by 16% to 32%, helping prevent muscle atrophy and reduce inflammation^[57, 58]^.Engaging in 1 hour of daily physical activity (e.g., leisure sports/housework) can reduce sarcopenia risk by 40% and decrease fall risk^[59]^.Strategy Adjustment: Given the trend of sarcopenia occurring at younger ages, the focus of prevention and control needs to shift forward to middle age (45-55 years). Implementing a tripartite strategy of metabolic management, nutritional optimization, and strength training is essential to disrupt the link between sarcopenia and fall risk.(3)Other Measures:Strategies to reduce systemic inflammation, clear senescent cells, or ameliorate mitochondrial dysfunction are beneficial for improving sarcopenia^[60]^.Myostatin inhibitors, zinc, catechins, soy isoflavones, and ursolic acid show positive effects on sarcopenia, although further research is needed for confirmation^[50]^.Emerging therapies such as whole-body vibration training, blood flow restriction training, and neuromuscular electrical stimulation show promise for sarcopenic individuals with limited mobility^[57]^.

The innovation of this study lies in: leveraging a large-scale longitudinal cohort (9-year follow-up, robust population representativeness) to pioneer focus on middle-aged and older adults, with particular emphasis on the sarcopenia-fall risk association in the younger elderly. It directly quantifies fall risk gradients across sarcopenia severity-stratified subgroups, transcending prior research confined to demographic subgroup risks. Employing RCS curves challenged traditional linear risk models by revealing a nonlinear dose-response relationship (risk gradient rising from Possible → Confirmed → Severe stages) and identified dual risk-rising periods and intervention windows, proposing the concept of “dual clinical intervention windows”.

Limitations of this study include the small sample size of severe sarcopenia, potentially limiting analytical power and masking true risk, necessitating validation with larger samples and expanded research on this subgroup. Secondly, the dynamic changes in sarcopenia status were not considered; future studies should incorporate multi-time-point data analysis to construct dynamic cohorts.

## 4 Conclusion

In conclusion, this study confirms a dose-response relationship between sarcopenia severity and fall risk. Sarcopenia is an independent graded risk factor for falls, with age 70 as a critical risk inflection point. Establishing a graded intervention system is essential, focusing on high-risk groups such as males, rural residents, smokers, drinkers, and those with chronic diseases. The intervention window needs to be shifted forward to middle age (45-55 years). Furthermore, interdisciplinary integration for prevention and control is crucial to establish a comprehensive sarcopenia-fall prevention system, reduce fall risk, and promote healthy aging.

## Acknowledgements

This research did not receive any specific grant from funding agencies in thepublic, commercial, or not-for-profit sectors.The authors wish to acknowledge all the participants who contributed to this study. Special thanks go to Dr. Shan Zheng for his mentorship throughout the research design and manuscript preparation.All acknowledged individuals have consented to be named in this publication.

## Data Availability Statement

The data that support the findings of this study are from the China Health and Retirement Longitudinal Study (CHARLS) database. These data are publicly available. Researchers can apply for access to the data via the official CHARLS website (http://charls.pku.edu.cn).

## Author Approval

All authors have read and approved the final manuscript.

## Authors’ Contributions

**Xia Yang:** Conceptualization, Methodology, Formal analysis, Writing – original draft, Writing – review & editing.

**Xue Wang:** Data curation, Investigation, Validation.

**Aiqin Zhao:** Software, Visualization.

**Haiye Wang:** Resources, Project administration.

**Shan Zheng:** Supervision, Writing – review & editing.

## Abbreviations

(CHARLS): China Health and Retirement Longitudinal Study
(RCS): Restricted Cubic Spline
(AWGS 2019): 2019 Asian Working Group for Sarcopenia
(SPPB): Short Physical Performance Battery
(GS): Gait speed
(DXA): Dual-energy X-ray absorptiometry
(BIA): Bioelectrical impedance analysis
(ASM): Appendicular skeletal muscle mass
(ASMI): Appendicular skeletal muscle mass index

## References

[1] SUN Min, WANG Ya, DING Zuoling. Establishment of a fall risk prediction model for elderly patients with sarcopenia based on the SMOTE algorithm [J]. Journal of Nursing Administration, 2024, 24(10):899–903.10.3969/j.issn.1671-315x.2024.10.014

[2] Cruz-Jentofta J, Baeyens J P, Bauer J M, et al. Sarcopenia: European consensus on definition and diagnosis [J]. Age and Ageing, 2010, 39(4):412–423.10.1093/ageing/afq034

[3] YUAN Shuai, Larsson S C. Epidemiology of sarcopenia: Prevalence, risk factors, and consequences [J]. Metabolism, 2023, 144: 155533.10.1016/j.metabol.2023.155533

[4] ZHENG Youle, FENG Jin, YU Yixin, et al. Advances in sarcopenia: mechanisms, therapeutic targets, and intervention strategies [J]. Archives of Pharmacal Research, 2024, 47(4): 301–324.10.1007/s12272-024-01493-2

[5] Petermann-Rocha F, Balntzi V, Gray S R, et al. Global prevalence of sarcopenia and severe sarcopenia: a systematic review and meta-analysis [J]. Journal of Cachexia, Sarcopenia andMuscle, 2021, 13(1): 86–99.10.1002/jcsm.12783

[6] YANG Menyu, YANG Yifang, WU Tong, et al. A nested case-control study on the effect of sarcopenia on mild cognitive impairment using the CHARLS database [J]. Geriatric Nursing, 2025, 61: 568–573.10.1016/j.gerinurse.2024.12.019

[7] GAO Ke, CAO Lifei, MA Wenzhuo, et al. Association between sarcopenia and cardiovascular disease among middle-aged and older adults: Findings from the China health and retirement longitudinal study [J]. EClinicalMedicine, 2022, 44: 101264.10.1016/j.eclinm.2021.101264

[8] Veronese N, Smith L, Barbagallo M, et al. Sarcopenia and fall-related injury among older adults in five low- and middle-income countries [J]. Exp Gerontol, 2021, 147: 111262.10.1016/j.exger.2021.111262

[9] ZHU Feiyun, YANG Ying, PAN Mengshan. Association between sarcopenia and all-cause mortality: a study based on CHARLS data from 2011 to 2020 [J]. Chinese Journal of Disease Control & Prevention, 2024, 28(10): 1170–1175.10.16462/j.cnki.zhjbkz.2024.10.009

[10] ZHANG Tiantian, FENG Zhiqiang, WANG Wanchen, et al. Study on the status and influencing factors of falls among Chinese older adults [J]. Chinese Journal of Disease Control & Prevention, 2022, 26(5): 502–507.10.16462/j.nki.zhjbkz.2022.05.002

[11] WANG Yilin, HUANG YaLian, CHEN Xiaoyan. The relationship between low handgrip strength with or without asymmetry and fall risk among middle-aged and older males in China: evidence from the China Health and Retirement Longitudinal Study [J]. Postgraduate Medical Journal, 2023, 99(1178): 1246-52..10.1093/postmj/qgad085

[12] LU Qiaochu, WANG Kang, ZHANG Luwen. Multimorbidity and fall risk among middle-aged and older adults in China: evidence from the CHARLS [J]. Journal of Applied Clinical Medicine, 2024, 40(13): 1851–1858.10.3969/j.issn.1006-5725.2024.13.015

[13] XIAO Xun, LI Ling, YANG Huijuan, et al. Analysis of the incidence of falls and related factors in elderly patients based on comprehensive geriatric assessment [J]. Aging Medicine, 2023, 6(3): 245–253.10.1002/agm2.12265

[14] Milisen K, Coussement J, Arnout H, et al. Feasibility of implementing a practice guideline for fall prevention on geriatric wards: A multicentre study [J]. International Journal of Nursing Studies, 2013, 50(4): 495–507.10.1016/j.ijnurstu.2012.09.020

[15] WANG Qianqian, ZHANG Yanzhuo, WU Cheng’ai. Analysis of risk factors for falls among middle-aged and older adults in China: based on the China Health and Retirement Longitudinal Study (CHARLS) data [J]. Chinese Journal of Gerontology, 2019, 39(15): 3794–3798.10.3969/j.issn.1005-9202.2019.15.061

[16] LIU Hao, HOU Yunfei, LI Hu, et al. Influencing factors of weak grip strength and fall: a study based on the China Health and Retirement Longitudinal Study (CHARLS) [J]. BMC Public Health, 2022, 22(1):2337..10.1186/s12889-022-14753-x

[17] Wilkinson A, Meikle N, Law P, et al. How older adults and their informal carers prevent falls: An integrative review of the literature [J]. International Journal of Nursing Studies, 2018, 82: 13–19.10.1016/j.ijnurstu.2018.03.002

[18] Chen L K, Woo J, AssantachaiS P, et al. Asian Working Group for Sarcopenia: 2019 Consensus Update on Sarcopenia Diagnosis and Treatment [J]. J Am Med Dir Assoc, 2020, 21(3): 300–7.e2.10.1016/j.jamda.2019.12.012

[19] LIU Haixia, ZHOU Ping, ZHANG Yina. Diagnosis and treatment of sarcopenia [J]. Chinese Journal of Osteoporosis and Bone Mineral Research, 2021, 14(4): 434–440.10.3969/j.issn.1674-2591.2021.04.015

[20] TANG Huan, LI Juan, YU Huan, et al. Research progress on muscle mass assessment tools for patients with sarcopenia [J]. Chinese Journal of Osteoporosis, 2023, 29(1):129–133.10.3969/j.issn.1006-7108.2023.01.025

[21] Curcio F, YANG Chunhua, YE Tengfei. Association between sarcopenia and falls in Chinese older adults:Findings from the China health and retirement longitudinal study [J]. PLoS One, 2025, 20(6): e0326193.10.1371/journal.pone.032619

[22] Zhang Xiaonin M, YE Dongmei, DOU Qingli, et al. Sarcopenia, Depressive Symptoms, and Fall Risk: Insights from a National Cohort Study in the Chinese Population [J]. Risk Management and Healthcare Policy, 2025, Volume 18: 593–603..10.2147/rmhp.S497087

[23] Marzettie, Calvanir, Coelho-Júnior H J, et al. Mitochondrial Quantity and Quality in Age-Related Sarcopenia [J]. International Journal of Molecular Sciences, 2024, 25(4).10.3390/ijms25042052

[24] Mcphee J S, Cameron J, Maden-Wilkinson T, et al. The Contributions of Fiber Atrophy, Fiber Loss, In Situ Specific Force, and Voluntary Activation to Weakness in Sarcopenia [J]. The Journals of Gerontology: Series A, 2018, 73(10): 1287–1294.10.1093/gerona/gly040

[25] LUO Xiaoqin, WANG Jin, JU Qingqing, et al. Molecular mechanisms and potential interventions during aging-associated sarcopenia [J]. Mech Ageing Dev, 2025, 223: 112020.10.1016/j.mad.2024.112020

[26] JI Weixiu. The role of skeletal muscle satellite cell-mediated muscle regeneration in the treatment of age-related sarcopenia [J]. Progress in Biochemistry and Biophysics, 2025: 2–18.10.16476/j.pibb.2025.0032

[27] Keller K, Engelhardt M. Strength and muscle mass loss with aging process. Age and strength loss [J]. Muscles Ligaments Tendons J, 2013, 3(4): 346–350. PMCID:PMC3940510

[28] Larsson L, Degens H, LI M, et al. Sarcopenia: Aging-Related Loss of Muscle Mass and Function [J]. Physiological Reviews, 2019, 99(1): 427–511.10.1152/physrev.00061.2017

[29] Dowling P, Gargan S, Swandulla D, et al. Fiber-Type Shifting in Sarcopenia of Old Age: Proteomic Profiling of the Contractile Apparatus of Skeletal Muscles [J]. International Journal of Molecular Sciences, 2023, 24(3).10.3390/ijms24032415

[30] LIU Dequan, WANG Shijin, LIU Shuang, et al. Frontiers in sarcopenia: Advancements in diagnostics,molecular mechanisms, and therapeutic strategies [J]. Mol Aspects Med, 2024, 97: 101270.10.1016/j.mam.2024.101270

[31] Nues-Pinto M, Bandeirera DE Mello R G, Pinto M N, et al. Sarcopenia and the biological determinants of aging: A narrative review from a geroscience perspective [J]. Ageing Res Rev, 2025, 103: 102587.10.1016/j.arr.2024.102587

[32] Keller K. Sarcopenia [J]. Wiener Medizinische Wochenschrift, 2018, 169(7-8): 157–172.10.1007/s10354-018-0618-2

[33] Lamboley C R, Wyckelsma V L, Dutka T L, et al. Contractile properties and sarcoplasmic reticulum calcium content in type I and type II skeletal muscle fibres in active aged humans [J]. The Journal of Physiology, 2015, 593(11): 2499–514.10.1113/jp270179

[34] Jun L, Robinson M, Geetha T, et al. Prevalence and Mechanisms of Skeletal Muscle Atrophy in Metabolic Conditions [J]. International Journal of Molecular Sciences, 2023, 24(3).10.3390/ijms24032973

[35] Kemmler W, Kohl M, FRöhlich M, et al. Effects of High-Intensity Resistance Training on Osteopenia and Sarcopenia Parameters in Older Men with Osteosarcopenia—One-Year Results of the Randomized Controlled Franconian Osteopenia and Sarcopenia Trial (FrOST) [J]. Journal of Bone and Mineral Research, 2020, 35(9): 1634–1644.10.1002/jbmr.4027

[36] WU Xin, LI Xue, XU Meihong, et al. Sarcopenia prevalence and associated factors among older Chinese population: Findings from the China Health and Retirement Longitudinal Study [J]. PLoS One, 2021, 16(3): e0247617.10.1371/journal.pone.0247617

[37] Marusic U, Narici M, Simunic B, et al. Nonuniform loss of muscle strength and atrophy during bed rest: a systematic review [J]. Journal of Applied Physiology, 2021, 131(1): 194–206.10.1152/japplphysiol.00363.2020

[38] LIANG Haodong, ZHANG Zijie, LAI Haitian, et al. Prevalence and risk factors for falls among older Chinese adults in the community: findings from the CLHLS study [J]. Braz J Med Biol Res, 2024, 57: e13469.10.1590/1414-431X2024e13469

[39] Lee A, Mcarthur C, Ioannidis G, et al. Associations between Osteosarcopenia and Falls, Fractures, and Frailty in Older Adults: Results From the Canadian Longitudinal Study on Aging (CLSA) [J]. Journal of the American Medical Directors Association, 2024, 25(1): 167–76.e6.10.1016/j.jamda.2023.09.027

[40] Vermeulen A. Ageing, hormones, body composition, metabolic effects [J]. World Journal of Urology, 2002, 20(1): 23–27.10.1007/s00345-002-0257-4

[41] LIM Jaeyoung, Frontera W R. Skeletal muscle aging and sarcopenia: Perspectives from mechanical studies of single permeabilized muscle fibers [J]. Journal of Biomechanics, 2023, 152.10.1016/j.jbiomech.2023.111559

[42] Sundberg C W, Hunter S K, Trappe S W, et al. Effects of elevated H+ and Pi on the contractile mechanics of skeletal muscle fibres from young and old men: implications for muscle fatigue in humans [J]. The Journal of Physiology, 2018, 596(17): 3993–4015.10.1113/jp276018

[43] JIANG Qian, DUAN Dechong, ZHANG Xiaodan. Correlation between sarcopenia and balance function in older women [J]. Chinese Journal of Rehabilitation Theory and Practice, 2020, 26(7): 842–846.10.3969/j.issn.1006-9771.2020.07.020

[44] Geraci A, Calvani R, FERRI E, et al. Sarcopenia and Menopause: The Role of Estradiol [J]. Front Endocrinol (Lausanne), 2021, 12: 682012.10.3389/fendo.2021.682012

[45] La Colla A, Pronsato L, Milanesi L, et al. 17β-Estradiol and testosterone in sarcopenia: Role of satellite cells [J]. Ageing Research Reviews, 2015, 24: 166–77.10.1016/j.arr.2015.07.011

[46] XU Qingmei, OU Xuemei, LI Jinfeng. The risk of falls among the aging population: A systematic review and meta-analysis [J]. Front Public Health, 2022, 10: 902599.10.3389/fpubh.2022.902599

[47] Chien S Y, Wang T H, Tzeng Y L, et al. Time Allocation to Physical Activity and Sedentary Behaviour and Its Impact on Sarcopenia Risk: A Systematic Review and Meta-Analysis [J]. Journal of Advanced Nursing, 2025, 81(10): 6250–60.10.1111/jan.16781

[48] Abiri B, Vafa M. Vitamin D and Muscle Sarcopenia in Aging [J]. Methods Mol Biol, 2020, 2138: 29–47.10.1007/978-1-0716-0471-7_2

[49] Burton L A, Sumukadas D. Optimal management of sarcopenia [J]. Clin Interv Aging, 2010, 5: 217–28.10.2147/cia.s11473

[50] Sakuma K, Yamaguchi A. Recent advances in pharmacological, hormonal, and nutritional intervention for sarcopenia [J]. Pflügers Archiv - European Journal of Physiology, 2017, 470(3): 449–460.10.1007/s00424-017-2077-9

[51] Papadopoulou S K. Sarcopenia: A Contemporary Health Problem among Older Adult Populations [J]. Nutrients, 2020, 12(5).10.3390/nu12051293

[52] WANG Yizhe, ZHAO Guoyang, YANG Jiaru. Mechanisms linking iron homeostasis dysregulation to sarcopenia [J]. Chinese Journal of Osteoporosis and Bone Mineral Research, 2023, 16(6): 603–608.10.3969/j.issn.1674-2591.2023.06.011

[53] WANG Dingkun, LI Yihao, GUO Xiaoming. Depression and sarcopenia-related traits: A Mendelian randomization study [J]. World Journal of Psychiatry, 2023, 13(11): 929–936.10.5498/wjp.v13.i11.929

[54] JIANG Yiwen, XU Bingqing, ZHANG Kaiyu, et al. The association of lipid metabolism and sarcopenia among older patients: a cross-sectional study [J]. Scientific Reports, 2023, 13(1).10.1038/s41598-023-44704-4

[55] Surakka J, AunolA S, Nordblad T et al. Feasibility of power-type strength training for middle aged men and women: self perception, musculoskeletal symptoms, and injury rates [J]. Br J Sports Med, 2003, 37(2): 131–136.10.1136/bjsm.37.2.131

[56] LU Linqian, MAO Lin, FENG Yuwei, et al. Effects of different exercise training modes on muscle strength and physical performance in older people with sarcopenia: a systematic review and meta-analysis [J]. BMC Geriatr, 2021, 21(1): 708.10.1186/s12877-021-02642-8

[57] HU Jiawen, Wang Y, iwen JI Xioajian, et al. Non-Pharmacological Strategies for Managing Sarcopenia in Chronic Diseases [J]. Clinical Interventions in Aging, 2024, Volume 19: 827–841.10.2147/cia.S455736

[58] Smeuninx B, Elhassan Y S, Sapey E, et al. A single bout of prior resistance exercise attenuates muscle atrophy and declines in myofibrillar protein synthesis during bed-rest in older men [J]. The Journal of Physiology, 2023, 603(1): 87–105.10.1113/jp285130

[59] Landi F, Liperoti R, Fusco D, et al. Prevalence and Risk Factors of Sarcopenia Among Nursing Home Older Residents [J]. The Journals of Gerontology Series A: Biological Sciences and Medical Sciences, 2011, 67A(1): 48-55.10.1093/gerona/glr035

[60] Jimenez-Gutierrez G E, Martínez-Gómez L E, Martínez-Armenta C, et al. Molecular Mechanisms of Inflammation in Sarcopenia: Diagnosis and Therapeutic Update [J]. Cells, 2022, 11(15).10.3390/cells11152359

